# Detection of Patients at Risk of Enterobacteriaceae Infection Using Graph Neural Networks: a Retrospective Study

**DOI:** 10.1101/2023.06.01.23290386

**Authors:** Racha Gouareb, Alban Bornet, Dimitrios Proios, Sónia Gonçalves Pereira, Douglas Teodoro

**Author notes:** Authors contributed equally (Racha Gouareb, Alban Bornet).

## Abstract

While Enterobacteriaceae bacteria are commonly found in healthy human gut, their colonisation of other body parts can potentially evolve into serious infections and health threats. We aim to design a graph-based machine learning model to assess risks of inpatient colonisation by multi-drug resistant (MDR) Enterobacteriaceae. The colonisation prediction problem was defined as a binary classification task, where the goal is to predict whether a patient is colonised by MDR Enterobacteriaceae in an undesirable body part during their hospital stay. To capture topological features, interactions among patients and healthcare workers were modelled using a graph structure, where patients are described by nodes and their interactions by edges. Then, a graph neural network (GNN) model was trained to learn colonisation patterns from the patient network enriched with clinical and spatiotemporal features. The GNN model predicts colonisation risk with an AUROC of 0.93 (95% CI: 0.92-0.94), 7% above a logistic regression baseline (0.86 [0.85-0.87]). Comparing different graph topologies, the configuration that considers only in-ward edges (0.93 [0.92-0.94]) outperforms the configurations that include only out-ward edges (0.86 [0.85-0.87]) and both edges (0.90 [0.89-0.91]). For the top-3 most prevalent MDR Enterobacteriaceae, the AUROC varies from 0.92 (0.90-0.93) for *Escherichia coli* up to 0.95 (0.92-0.98) for *Enterobacter cloacae*, using the GNN – in-ward model. Topological features via graph modelling improves the performance of machine learning models for Enterobacteriaceae colonisation prediction. GNNs could be used to support infection prevention and control programmes to detect patients at risk of colonisation by MDR Enterobacteriaceae and other bacteria families.

## Introduction

Healthcare-associated infection (HAI) is a severe health problem for patients, health professionals and visitors in a healthcare facility^1,2^. The World Health Organization estimates that one in every ten patients develops an HAI^3^ and, in US hospitals alone, the Centers for Disease Control estimate that HAIs account for 1.7 million infections and 99’000 associated deaths each year^4^. Among these infections, more than one third are caused by Enterobacteriaceae^5^, a family of bacteria that includes the most prevalent human pathogenic species and the leading causes of nosocomial infections, such as *Escherichia coli, Salmonella enterica*, and *Klebsiella pneumoniae*. Given that these infections are acquired in environments under high antimicrobial pressure, they are often caused by antimicrobial resistant (AMR) and multidrug resistant (MDR) bacteria. MDR Enterobacteriaceae infections have augmented drastically over the last two decades, especially with the rise of carbapenemase-producing Enterobacteriaceae^6^. These pathogens are able to resist not only to the action of all available beta-lactams (except aztreonam), but also to other available antimicrobial classes like fluoroquinolones and aminoglycosides, leaving physicians with few treatment options^7^. This leads to more expensive treatments, longer hospital stays, increased risks of complication, and higher risks of death^8^.

The continuous rise of these pathogens in healthcare settings is multifactorial, with their ability to spread and persist in the environment and asymptomatically in patients and healthcare workers accounting as main contributors^9^. The risk of colonisation, subsequent infection and mortality due to Enterobacteriaceae increases exponentially with age, health history, and length of hospital stay^10^. Colonisation can be defined as the asymptomatic presence of a pathogen in the human body. It is not only the first step towards an overt disease of the colonised patient, with more or less severity, but also one of the main contributors to infection outbreaks in healthcare settings^11^. Indeed, some studies showed that between 36% and 39% of patients colonised by AMR Enterobacteriaceae develop a subsequent infection^12,13^. Asymptomatic infections, specially by MDR bacteria, pose also a prominent public health issue as the pathogen that the colonised patient carries can inadvertently be transmitted to other patients, which can become only colonised or, more concerningly, symptomatically infected, with increased risks of complications and even death^6^. Infection prevention and control (IPC) programs provides critical measures for preventing disease transmission in healthcare settings, with the potential to lower HAI rates by at least 30%, being sometimes the only solution to prevent and avoid these MDR colonisations and infections.

Leveraging the availability of large-scale healthcare data^14–16^, routinely collected and stored in electronic health records (EHRs), machine learning models have been proposed for early detection of patients at risk of infection and to support IPC programs^17–21^. Classic machine learning methods, such as decision trees and random forest, have demonstrated good performance to predict patients at risk of HAI^22–25^. For methicillin-resistant *Staphylococcus aureus*^22^ and *Clostridioides difficile*^25^, these algorithms were shown to provide warnings as early as five days before diagnosis. Machine learning methods for colonisation prediction was also explored in very recent studies^26–28^. Tree-based machine learning methods, such as decision trees, random forest and extreme gradient boosting, achieved sensitivity and specificity above 80% for detecting MDR species from different pathogenic families^27^, while the use of spatiotemporal features to identify patients colonised by vancomycin-resistant Enterococcus resulted in area under the receiver operating characteristic curve (AUROC) performance above 88%^26^.

While classic machine learning models and hand-crafted features might show effective results in limited use cases, they often fail to generalize to large-scale and longitudinal EHR data^29,30^. Another limitation of previous approaches for Enterobacteriaceae colonisation prediction is that key interactions between patients and healthcare workers are neglected, hindering their application to complex care networks. To address these gaps, we propose a deep-learning approach based on a graph neural network (GNN) architecture^31^. This approach aims to incorporate interactions between patients and healthcare workers, inside and outside the wards, as well as other clinical and spatiotemporal features, to predict risks of Enterobacteriaceae colonisation for inpatients. Our models were trained and evaluated using the Medical Information Mart for Intensive Care (MIMIC-III) dataset^32^ and compared with classic machine learning baselines. Interestingly, the GNN models provide stronger predictive performance for early detection of AMR and MDR Enterobacteriaceae, compared to models trained on data without the patient network information. Our main contributions can be summarized as follows:

- We propose a graph-based colonisation model that considers spatiotemporal features in addition demographic and clinical condition. To avoid adding biases to the model due to information leakage, we deliberately did not use antimicrobial information.
- We design a new machine learning architecture for colonisation prediction using a GNN architecture that learns transmission network patterns from spatiotemporal and patient data. Different network configurations and transmission paths were proposed and evaluated.
- We evaluate our model against classic state-of-the-art machine learning baselines and show that it achieves superior performance, both for the original dataset and for an alternate version of the dataset that is free of class imbalance. We also conducted an explainability study to demonstrate the capacity of the model to automatically identify features associated with colonisation risk factors.
- There have been many studies investigating HAI prediction. To the best of our knowledge, this is the first attempt to explore the problem of predicting risks of AMR and MDR Enterobacteriaceae colonisation for undesirable body parts using graph models and provide data-driven hypothesis for transmission.

## Methods

### Study design and data sources

To train and evaluate our colonisation risk prediction models, we used laboratory, clinical, and administrative data from patients who stayed in critical care units of the Beth Israel Deaconess Medical Center (Massachusetts, USA). These data were recorded between 2001 and 2012 and made publicly available through the MIMIC-III dataset^32^. MIMIC-III is a freely available and deidentified healthcare dataset that consists of 26 tables and includes static and dynamic patient information, such as demographics, medical history and records, clinical measures, laboratory tests, and interventions. The database contains data from 46’520 unique patients aged 16 years or older and associated to 58’976 admissions. Patients can be admitted to the hospital more than once and moved between 50 different wards and seven care units during their stays. Additionally, activities from 7’567 unique healthcare workers - a nurse or a medical doctor - are recorded.

In the MIMIC-III dataset, we observed that 17% of inpatients had a positive result for Enterobacteriaceae screen. In total, 14 different bacterial species of the Enterobacteriaceae family were found from a total of 30 unique specimen types collected from inpatients. Figure 1 shows their distribution for different sample types (Figure 1-left) and different resistant profiles (Figure 1-right). *E. coli* was the most frequently found positive culture (50%), while *Citrobacter amalonaticus* or *Salmonella enterica* (not shown), were rarely found. A bacterial isolate was considered AMR if showing resistance to at least one agent in only one or two antimicrobial categories, and MDR if it was resistant to at least one agent in three or more antimicrobial categories. Otherwise, it was classified as antimicrobial susceptible (AMS). As shown in Figure 1, *C. koseri, E. coli*, and *K. pneumonia* were the species with highest levels of resistance (>50%), the latter two showing MDR profiles in more than 25% of cases.

**Figure 1:**
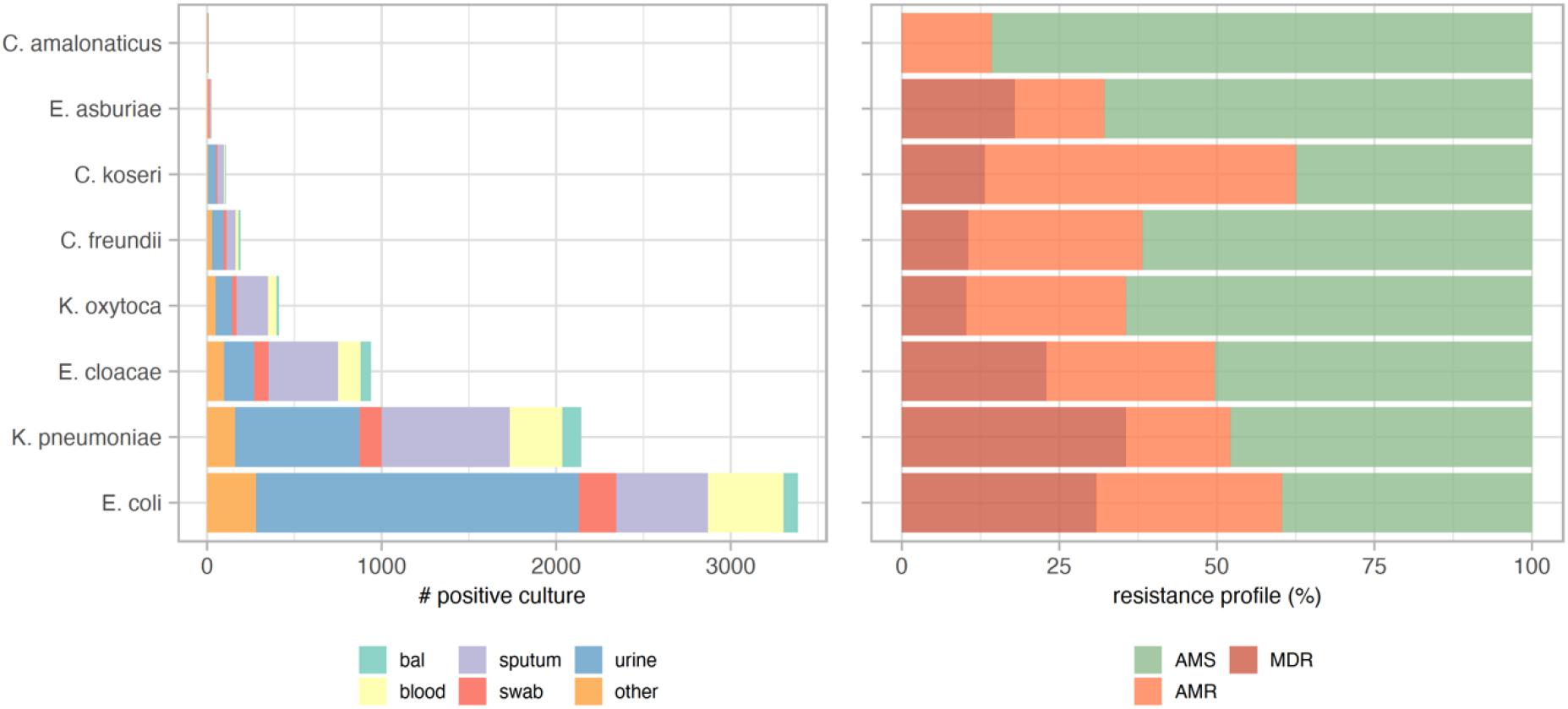
Frequency of positive culture and resistant profile for each Enterobacteriaceae family. Only species with more than 5 positive cultures are shown. Bal: bronchoalveolar lavage.

The training and evaluation dataset used in this study was created using the cohort selection criteria described in Figure 2. The *Microbiology Events* table from MIMIC-III was used to detect positively colonised patients. The table contains bacterial identification and antimicrobial testing results, and consists of 631’726 events related to 46’520 patients. A list of Enterobacteriaceae species was selected using the National Center for Biotechnology Information terminology^33^ and used to select the microbiology events of patients colonised by Enterobacteriaceae. This first step resulted in 109’318 events related to 4’868 colonised patients. Then, a list of abnormal specimens (or uncommon body parts) where these species were found was identified by two clinical microbiologist experts and categorized into six specimen categories: blood, gastric-related, respiratory, skin, tissue, and urine. This list defined the set of positive colonisation events that were relevant to our study, i.e., presence of Enterobacteriaceae in abnormal body parts, resulting in 107’313 microbiology events and 4’838 colonised patients. Finally, the *Admissions* table provided information regarding every unique hospitalization for every patient in the database. The table was used to define the remaining non-colonised patients. Amongst all admitted patients, the ones that were not found in the filtered *Microbiology Events* table, in addition to those with Enterobacteriaceae in regular specimens (i.e., stool samples), were considered non-colonised. Lastly, the table *Transfers*, which contains patient location information and their transfers between wards, was used to assign patients to wards. The final study dataset contained 46’520 unique patients from 58’976 admissions, and a total of 274’316 patient-ward instances. If during a whole stay in a ward, there was no positive abnormal Enterobacteriaceae culture for a patient, the patient-ward instance was labelled as non-colonised; otherwise, as colonised. This resulted in 7’216 positive Enterobacteriaceae colonisations (2.6%) and 267’100 negative specimens (97.4%). The dataset was randomly divided into train (60%), dev (20%) and test (20%) sets to train the machine learning model parameters, optimize the hyper parameters and evaluate the performance, respectively. Each set contained around 2.5-3% of colonised patients.

**Figure 2:**
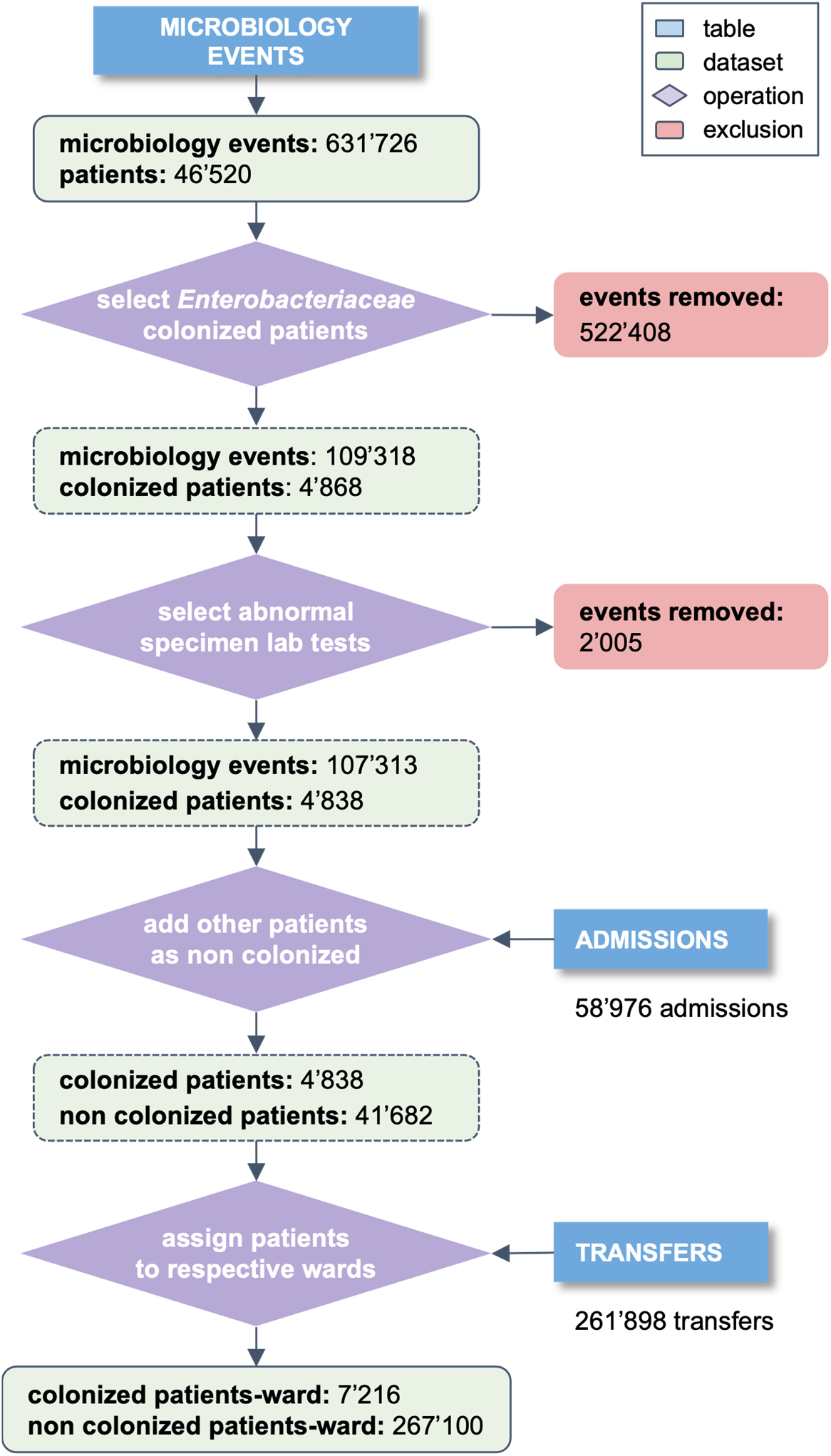
Cohort selection criteria. Starting from the *Microbiology Events* table of MIMIC-III, lab results were filtered for the existence of bacteria belonging to the Enterobacteriaceae family in unusual body parts to define colonised patients. *Admissions* and *Transfers* tables were used to complement the remaining patients and to label all patients.

### Feature selection and data pre-processing

The feature selection process was performed iteratively. The MIMIC-III dataset was first analysed to pre-identify the set of features we considered relevant to the colonisation risk prediction problem. Then, based on model’s performance computed on the dev set, less significant features, such as the death time and the discharge status of the patient, were eliminated. The final feature set can be grouped into two types: *i)* spatiotemporal features (current and previous ward, current and previous care unit, length of stay in each ward and in the hospital) and *ii)* patient features (gender and diagnosis at admission). To complement this set, we computed three new features from the data: the number of colonised patients, the total number of patients per ward, and the colonisation pressure^34^. The latter was calculated as the ratio of colonised and the total number of patients in a ward per day. Finally, the features were normalized using the *robust scaler* method of scikit-learn^35^, version 1.1.2. The statistics of the resulting dataset are shown in Table 1.

**Table 1:** Statistics of the cohort used for model training and evaluation.

|  | Non-colonised (n = 267'100) | Colonised (n = 7'216) |
| --- | --- | --- |
| Sex |  |  |
| female | 116'886 (43.8%) | 3'631 (50.3%) |
| male | 150'214 (56.2%) | 3'585 (49.7%) |
| Age (years) |  |  |
| 0-17 | 35'446 (13.3%) | 152 (2.1%) |
| 18-25 | 6'325 (2.4%) | 130 (1.8%) |
| 26-45 | 29'023 (10.9%) | 754 (10.4%) |
| 46-65 | 83'912 (31.4%) | 2'436 (33.7%) |
| 66-88 | 101'629 (38.0%) | 3'461 (48.0%) |
| ≥ 89 | 10'765 (4.0%) | 283 (4.0%) |
| Reason for admission |  |  |
| newborn | 34'162 (12.8%) | 122 (1.7%) |
| pneumonia | 6'736 (2.5%) | 140 (1.9%) |
| sepsis | 4'603 (1.7%) | 222 (3.1%) |
| coronary artery disease | 4'449 (1.7%) | 52 (0.7%) |
| congestive heart failure | 4'236 (1.6%) | 121 (1.7%) |
| chest pain | 3'905 (1.5%) | 75 (1.0%) |
| other | 209'009 (78.2%) | 6'484 (89.9%) |
| Resistance profile |  |  |
| AMS | – | 3288 (45.6%) |
| AMR | – | 3928 (54.4%) |
| MDR | – | 2099 (29.1%) |
| Length of stay (days) |  |  |
| 0-4 | 57'511 (21.5%) | 407 (5.6%) |
| 5-10 | 105'531 (39.5%) | 1'566 (21.7%) |
| 11-50 | 93'651 (35.1%) | 4'284 (59.4%) |
| 51-100 | 8'183 (3.1%) | 738 (10.2%) |
| ≥ 100 | 2'224 (0.8%) | 221 (3.1%) |
| Specimen |  |  |
| urine | – | 2'954 (40.9%) |
| sputum | – | 1'944 (26.9%) |
| blood | – | 934 (12.9%) |
| swab | – | 483 (6.7%) |
| bronchoalveolar lavage | – | 288 (4.1%) |
| other | – | 613 (8.5%) |
| Enterobacteriaceae species |  |  |
| *E. coli* | – | 3'385 (47.0%) |
| *K. pneumoniae* | – | 2'142 (29.7%) |
| *E. cloacae* | – | 939 (13.0%) |
| *K. oxytoca* | – | 410 (5.7%) |
| *C. freundii* | – | 191 (2.6%) |
| *C. koseri* | – | 107 (1.5%) |
| other | – | 42 (0.5%) |

### Colonisation network model

We propose a homogeneous graph to model interactions between patients and healthcare workers. A graph can be defined as *G =* (*V, E*), where *V = v*_1_, …, *v*_|*V*|_ denotes a set of nodes and *E* denotes a set of edges connecting pairs of nodes *v*_*i*_, *v*_*j*_ ∈ *V*. In our case, a node represents a patient and edges represent potential connections between them, either via contact with the same healthcare worker or via a common location within the hospital. As shown in Figure 3a, we considered three network configurations: *i) in-ward links* (left), where two patients are linked only if they stay in the same ward at the same time, *ii) out-ward links* (middle), where two patients are connected only if they are visited by the same healthcare worker on the same day, and *iii) all links* (right), where both ward and healthcare worker links are considered. Nodes represent a patient in a ward and their features are created using the selected feature set described in the previous section. When a patient is transferred, a new node is added to the graph with its corresponding new edges according to the different network configurations previously described (i.e., in-ward, out-ward and all links).

**Figure 3:**
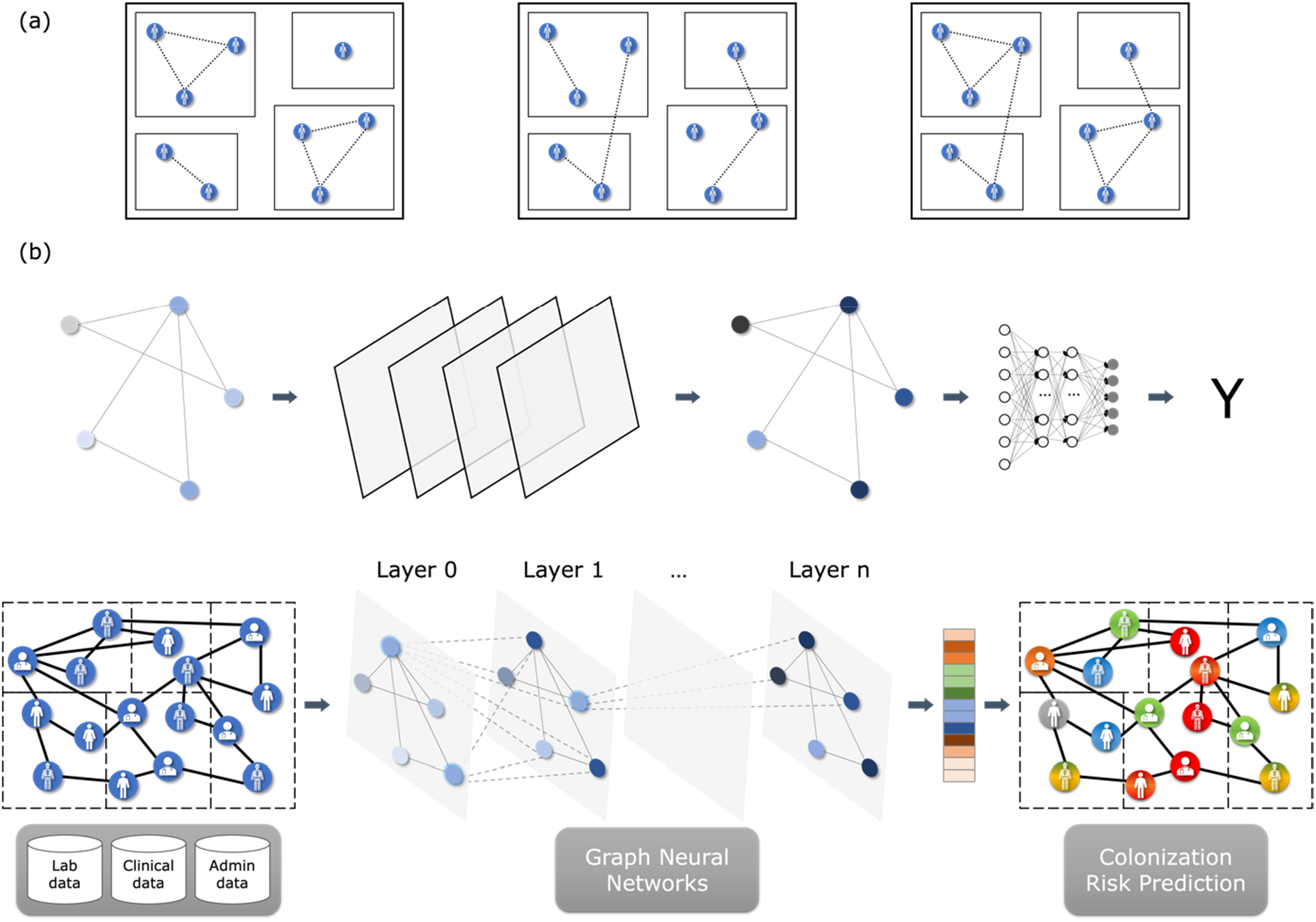
(a) Colonisation models. We constructed 3 different graphs, in which links were created between patients only if they were in the same ward (left), only if they were visited by the same healthcare worker (centre) or both (right). (b) Graph-based machine learning pipeline for colonisation risk prediction.

### Graph neural network architecture

An elegant deep learning architecture for modelling graph-like data structures is GNN^31,36–38^ and learn topological features, i.e., properties of the transmission network in our case. GNNs can learn complex relationships and interdependencies in graph-like data via optimizable transformations on attributes (nodes, edges, etc.) that preserve graph symmetries (i.e., permutation invariance/equivariance). Hence, in theory GNNs can make more informed predictions about entities in a network and their interactions, as compared to models that consider entities in isolation. To solve graph representation learning tasks, different GNN network architectures and algorithms have been proposed, such as graph convolutional network (GCN)^31^, graph attention networks^39^, and GraphSAGE^40^. These approaches use various graph feature aggregation and data sampling strategies to learn dense representations of graph components (i.e., nodes and edges), often called embeddings, that can be later used in downstream prediction tasks, such as node classification.

In our experiments, we used the GCN architecture, which employs convolutional aggregations to create graph features. GCNs are invariant to node permutations, which means that isomorphic graphs result in the same learned representation. In GCNs, node representations are learned based on neighbouring node features, which are propagated across the graph using the message passing algorithm^41^. At each layer of the GCN, every node of the graph is represented by a hidden state 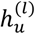, where *u* indexes the node and *l* the GCN layer that encodes the node features. An aggregation function *A* is used to send information from the immediate neighbourhood *v* ∈ *N*(*u*) to every node *u*. Finally, to update each node representation 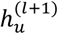 in the subsequent layer *l* + 1 of the network, an update function *U* is used, i.e., 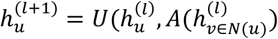. In our case, the aggregation function is the sum (the hidden states of neighbouring nodes are summed over the node dimension), and the update function is a multi-layer perceptron.

A high-level view of the graph-based prediction pipeline is shown in Figure 3. Using laboratory, clinical and administrative data, patient features at the ward level (i.e., a node represented a patient in a ward) were extracted and modelled in different network colonisation models (Figure 3a). The colonisation graph was fed to a two-layer GCN (*L =* 2), with embedding size set 32, followed by a sigmoid layer for classification (Figure 3b). A binary cross-entropy with logits loss was used to train the models. To account for data imbalance, the loss coming from the non-colonised class was weighted using the ratio of non-colonised patient to the total number of patients during training. The decision threshold was set to 0.9, i.e., a patient was classified as colonised if the inferred colonisation risk was over 0.9, and otherwise classified as non-colonised. We trained all models for 400 epochs, using the Adam optimizer. The number of layers, embedding size, number of epochs and decision threshold were tuned using the dev set.

### Statistical analysis

To evaluate the performance of the colonisation risk prediction models, standard binary classification metrics were computed: accuracy, macro-averaged precision, recall, F1-score, and AUROC. The GNN models were compared to classic machine learning baselines: k-nearest neighbours (kNN)^42^, logistic regression^43^, random forest^44^ and CatBoost^45^. Student t-test was used to compare model performance. Results were deemed statistically significant for *p*-value smaller than 0.05. Shapley values were used to measure the importance of each feature to the model’s predictions.

## Results

Results obtained with the different colonisation risk prediction models are presented in Table 2. In addition to the individual classic and graph-based models (Table 2A), we created three types of ensemble models (Table 2B): *i*) *ensemble - classic*, which combines the results of classic machine learning models; *ii*) *ensemble - GNN*, which combines the results of GNN models; and *iii*) *ensemble - all*, which combines the results of all models. For each ensemble type, we applied three voting strategies: *i*) unanimity vote [25], where a prediction is considered only if all model participants vote for the same class, and rejected otherwise; *ii*) majority vote, where a prediction is considered only if it reaches the majority amongst model participants, and rejected if there is an equality; *iii*) average probability, where the predicted class probabilities are averaged over all models to generate a prediction. Comparing the individual models, the GNN-based models show strong AUROC performance, all above 86% and outperform the classic machine learning models (*p*-value *<* 0.001) for this metric. Particularly, the GNN model that uses *in-ward* topology only (*GNN - in-ward*) achieves the highest AUROC (92.59% [95% confidence interval (CI): 91.67-93.51]), outperforming all individual classic models (*p*-value *<* 0.001). Within the graph-based models, the *out-ward* topology shows the weakest performance (86.20% [95% CI: 85.01-87.40]) followed by the *all-links* topology (89.58% [95% CI: 88.51-90.65]) (*p*-value *<* 0.001). These results suggest that network features enhance the predictive power of machine learning models for colonisation risk prediction, and that transmission patterns within the same ward are more useful features.

**Table 2:**
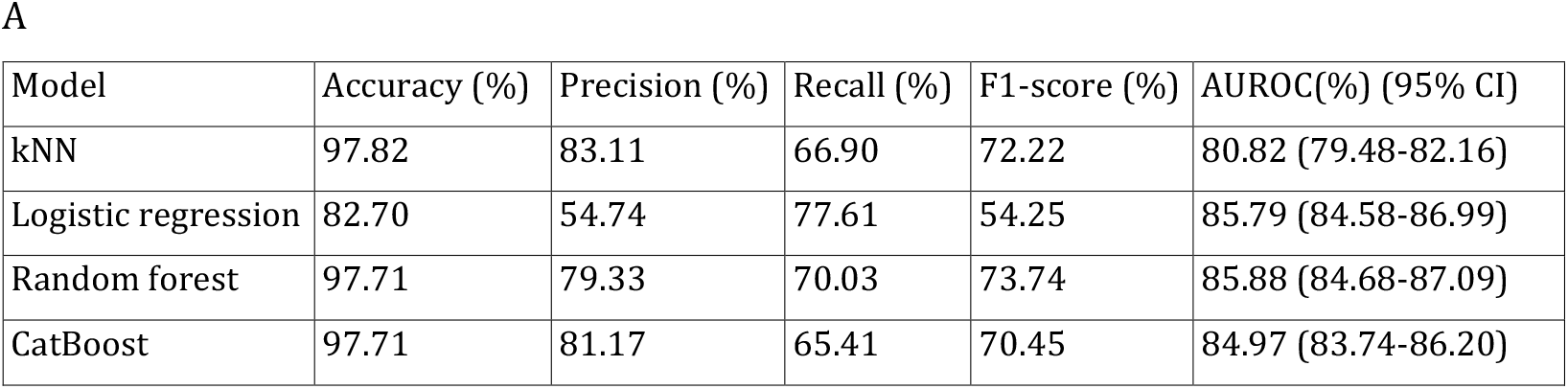
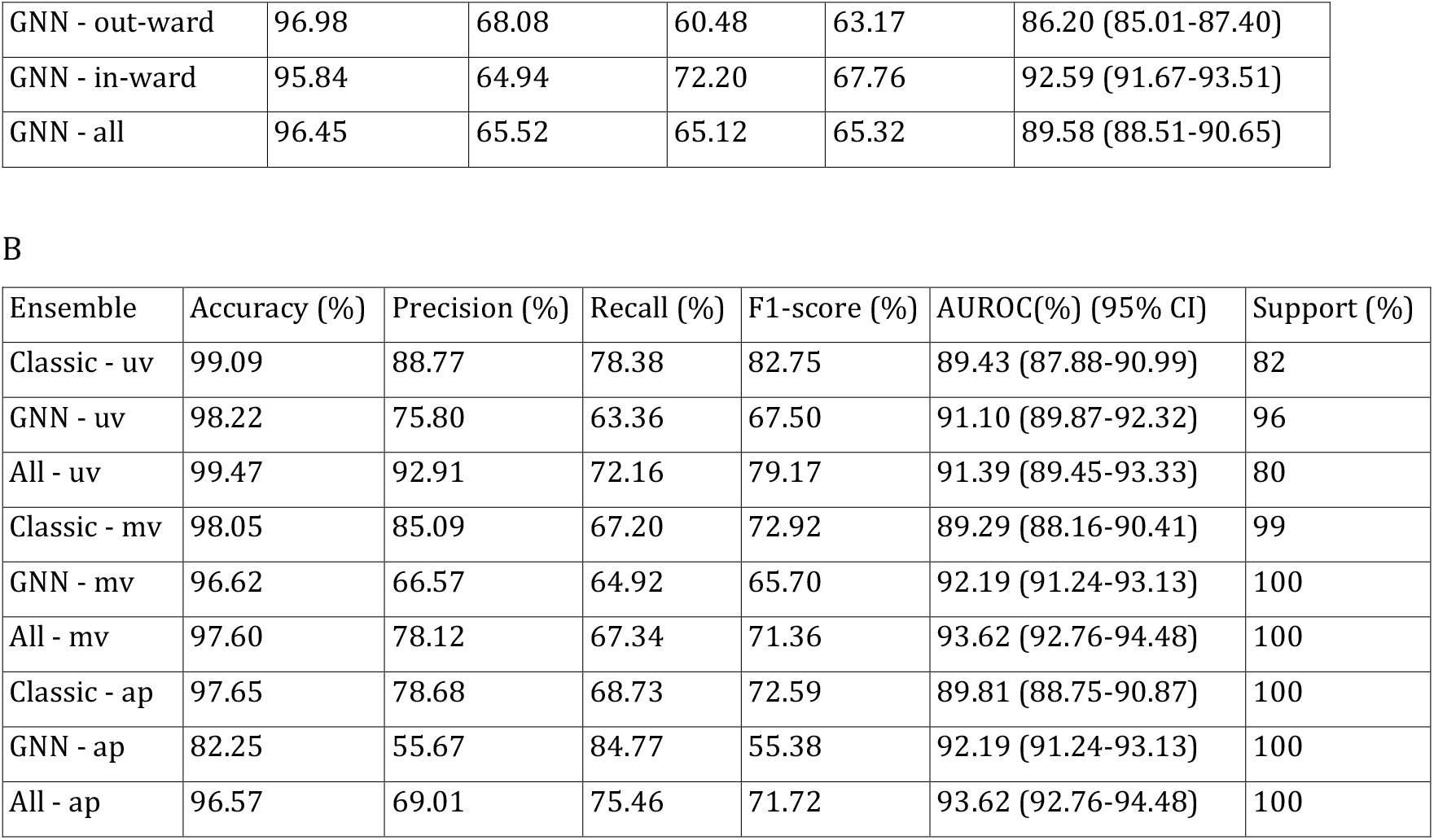
Performance of the individual colonisation prediction models (A) and of the ensemble models (B), based on three voting strategies: *i)* unanimity vote (uv), *ii)* majority vote (mv) and *iii)* average probability (ap). Precision, recall and F1-score are macro-averaged.

| Model | Accuracy (%) | Precision (%) | Recall (%) | F1-score (%) | AUROC(%) (95% CI) |
| --- | --- | --- | --- | --- | --- |
| kNN | 97.82 | 83.11 | 66.90 | 72.22 | 80.82 (79.48-82.16) |
| Logistic regression | 82.70 | 54.74 | 77.61 | 54.25 | 85.79 (84.58-86.99) |
| Random forest | 97.71 | 79.33 | 70.03 | 73.74 | 85.88 (84.68-87.09) |
| CatBoost | 97.71 | 81.17 | 65.41 | 70.45 | 84.97 (83.74-86.20) |
| GNN - out-ward | 96.98 | 68.08 | 60.48 | 63.17 | 86.20 (85.01-87.40) |
| GNN - in-ward | 95.84 | 64.94 | 72.20 | 67.76 | 92.59 (91.67-93.51) |
| GNN - all | 96.45 | 65.52 | 65.12 | 65.32 | 89.58 (88.51-90.65) |

B
| Ensemble | Accuracy (%) | Precision (%) | Recall (%) | F1-score (%) | AUROC(%) (95% CI) | Support (%) |
| --- | --- | --- | --- | --- | --- | --- |
| Classic - uv | 99.09 | 88.77 | 78.38 | 82.75 | 89.43 (87.88-90.99) | 82 |
| GNN - uv | 98.22 | 75.80 | 63.36 | 67.50 | 91.10 (89.87-92.32) | 96 |
| All - uv | 99.47 | 92.91 | 72.16 | 79.17 | 91.39 (89.45-93.33) | 80 |
| Classic - mv | 98.05 | 85.09 | 67.20 | 72.92 | 89.29 (88.16-90.41) | 99 |
| GNN - mv | 96.62 | 66.57 | 64.92 | 65.70 | 92.19 (91.24-93.13) | 100 |
| All - mv | 97.60 | 78.12 | 67.34 | 71.36 | 93.62 (92.76-94.48) | 100 |
| Classic - ap | 97.65 | 78.68 | 68.73 | 72.59 | 89.81 (88.75-90.87) | 100 |
| GNN - ap | 82.25 | 55.67 | 84.77 | 55.38 | 92.19 (91.24-93.13) | 100 |
| All - ap | 96.57 | 69.01 | 75.46 | 71.72 | 93.62 (92.76-94.48) | 100 |

For the metrics with a decision threshold, kNN achieves the highest accuracy, with a performance of 97.82% (*p*-value *<* 0.001). Apart from logistic regression (82.70%), all models achieve strong accuracy, with values of 96% or higher. Due to the large class imbalance, this strong performance is expected for the accuracy metric. It is particularly noteworthy though that the *ensemble - all* model, following the unanimity vote strategy, obtained an accuracy of 99.47% when classifying 80% of the test set (the remaining 20% were discarded due to the lack of convergence between the individual classifiers). For the macro-average metrics, random forest achieves the highest F1-score among the individual models, with a performance of 73.74% (*p*-value *<* 0.001), nearly 6% above the best GNN model. This is likely due to an overfitting of the decision threshold for GNNs (which was trained using the dev set). Ensemble models also improve significantly upon individual models for the macro-average metrics, with an F1-score of up to 82.75% for the *ensemble - classic* configuration following the unanimity vote strategy, while classifying 82% of the samples. Lastly, as shown by the macro-average metrics (precision, recall and F1-score), the ensemble models achieve a more balanced predictive performance between colonised and non-colonised patients, which together with a high accuracy may foster better practical applications (at the expense of a reduced assessment set).

### Stratified performance analysis for the GNN model

Figure 4 presents the AUROC performance of the best individual model - *GCN - in-ward* - stratified by species, specimen type, length of stay and resistance profile. The results show that the model provides consistent performance across different bacteria species, with AUROC above 89% for species that have at least 7 examples in the training dataset. The best performance is seen for *E. cloacae* (94.08% [95% CI: 91.70-96.45]) (939 examples in the training set) while the worse is for *C. amalonaticus* (95% CI: 79.60% [95% CI: 27.02-100.00]) (7 examples in the training set). Similarly, consistent performance is observed across specimens, with AUROC varying from 90.58% (95% CI: 87.85-93.31) for *blood* culture to 95.00% (95% CI: 93.53-96.47) for *sputum*. The results of Figure 4c show a decreasing trend in performance as patients stay longer in the hospital (R^2^=0.8347), with performance as high as 94.14% (95% CI: 90.99-97.28) for patients that stay 4 days or less and as low as 85.60% (95% CI: 78.62-92.59) for patients that stay more than 100 days. Lastly, the model achieves similar predictive performance for different resistance profiles with the lowest score of AUROC at 92.26% (95% CI: 90.90-93.63) for AMS Enterobacteriaceae and the highest score at 93.25% (95% CI: 91.58-94.92) for MDR Enterobacteriaceae.

**Figure 4:**
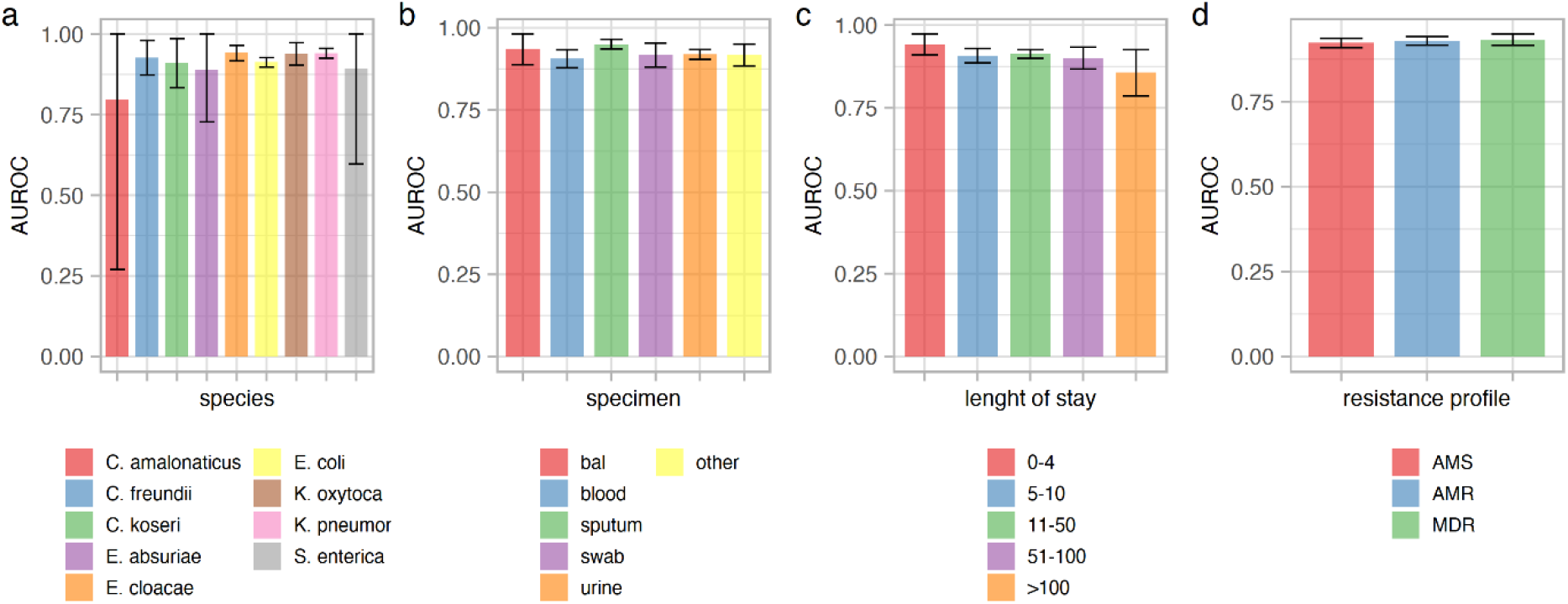
Performance results per species, specimen type, length of stay and resistance profile. bal: bronchoalveolar lavage; AMS: antimicrobial susceptible; AMR: antimicrobial resistant; MDR: multi-drug resistant.

### Predictive performance for AMR and MDR resistance profiles

The predictive performance according to AMS, AMR and MDR resistance profiles and for the three most frequent MDR Enterobacteriaceae species is shown in Figure 5. Similar to the overall case, the *GNN – in-ward* and *Ensemble – all* models show robust performance across the difference resistance profiles and species, outperforming all the respective individual and ensemble models. For the AMR and MDR resistance profiles, the *GNN – in-ward* model achieved an AUROC of 92.89% (95% CI: 92.13-93.65) and 93.25% (95% CI: 92.28-94.21) respectively, which were slightly outperformed by the *Ensemble – all* model (93.87% [95% CI: 93.10-94.64] and 94.33% [95% CI: 93.32-95.34], respectively). For the top-3 most prevalent MDR Enterobacteriaceae, the AUROC varied from 91.74% (95% CI: 90.25-93.23) for *E. coli* up to 95.16% (95% CI: 91.92-98.41) for *E. cloacae* using the *GNN – in-ward* model, and from 92.63% (95% CI: 90.99-94.28) for *E. coli* up to 96.33% (95% CI: 95.16-97.50) for *K. pneumoniae* using the *Ensemble – all* model. Results for the classic models are slightly less consistent, with logistic regression (AMR, MDR, MDR *E. coli* and MDR *E. cloacae*), random forest (MDR *K. pneumoniae*) and CatBoost (AMS) claiming the best performance depending on the test set strata.

**Figure 5:**
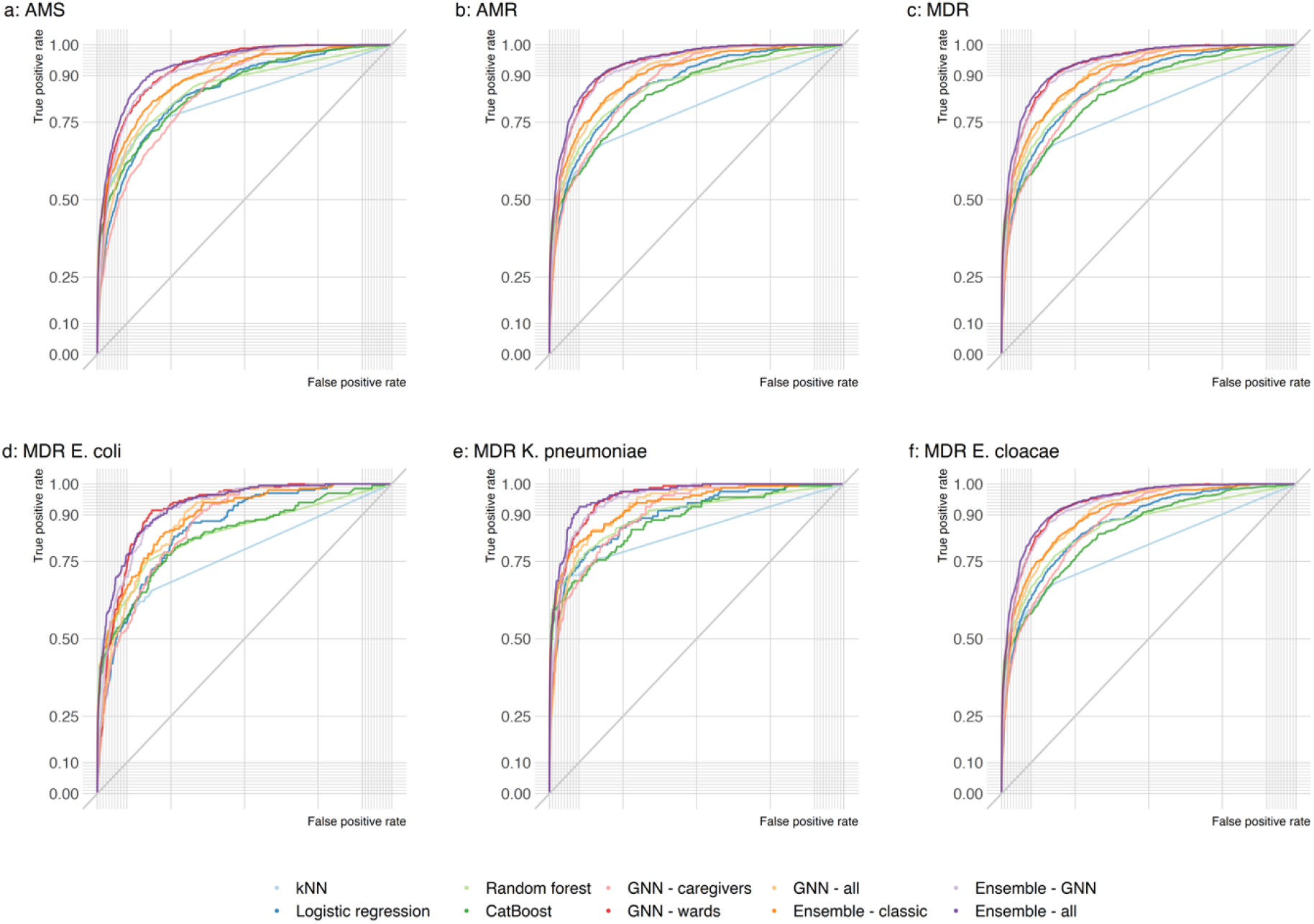
Model performance for antimicrobial susceptible (AMS), resistant (AMR) and multi-drug resistant (MDR) Enterobacteriaceae.

### Balanced dataset scenario

As the distribution of colonised and non-colonised patients in our dataset is highly imbalanced, we evaluated the proposed models on a balanced dataset, representing an optimal-case scenario for machine learning methods. In our experiments, the balanced dataset was generated by under-sampling the original database, resulting in 8658 samples for the training set and 2887 samples for the test set, including 1473 non-colonised (51%) and 1414 colonised (49%) examples. As shown in Table 3, the results of the balanced scenario followed a similar pattern as for the original dataset. Amongst individual models, the best performance in terms of AUROC was again achieved by the GNN models, more specifically, *GNN in-ward*, with an of AUROC 92.08% (95% CI: 91.03-93.12) (*p*-value *<* 0.001). Similar to the original data scenario, for the metrics with a decision threshold, the *GNN - in-ward* model reached the highest performance in the balanced setup, with an accuracy of 80.04% (*p*-value *<* 0.001) and an F1-score of 79.61% (*p*-value *<* 0.001), outperforming even the ensemble approach based on the average probability vote strategy.

**Table 3:** Performance of the colonisation prediction models using a balanced dataset. Precision, recall and F1-score are macro-averaged. The ensemble model was built based on the average probability strategy.

| Model | Accuracy (%) | Precision (%) | Recall (%) | F1-score (%) | AUROC (%) (95% CI) |
| --- | --- | --- | --- | --- | --- |
| kNN | 72.73 | 72.72 | 72.72 | 72.72 | 81.57 (80.00-83.13) |
| Logistic regression | 71.70 | 71.90 | 71.58 | 71.55 | 78.70 (77.03-80.36) |
| Random forest | 77.76 | 77.77 | 77.78 | 77.76 | 86.62 (85.27-87.97) |
| CatBoost | 78.38 | 78.42 | 78.42 | 78.38 | 86.06 (84.69-87.44) |
| GNN - out-ward | 71.14 | 79.73 | 71.68 | 69.22 | 85.76 (84.37-87.15) |
| GNN - in-ward | 80.04 | 83.67 | 80.38 | 79.61 | 92.08 (91.03-93.12) |
| GNN - all | 73.92 | 79.76 | 74.36 | 72.79 | 88.76 (87.52-90.01) |
| Ensemble - classic | 78.14 | 78.14 | 78.15 | 78.14 | 86.56 (85.21-87.91) |
| Ensemble - GNN | 65.01 | 79.06 | 65.71 | 60.77 | 91.44 (90.36-92.53) |
| Ensemble - all | 76.06 | 82.19 | 76.50 | 75.06 | 89.06 (87.83-90.28) |

### Feature impact on model predictions

To explain the importance and impact of the features used in our colonisation risk prediction models, we calculated their Shapley values using the SHAP method^46^. For simplicity, we used the results of the random forest model as the one with the highest F1-score in the original dataset. Figure 6a shows the importance of the top-11 features sorted by their predictive impact, with the most significant features on top and the least important ones at the bottom. Figure 6b shows the mean absolute value of every feature presented in Figure 6a, computed over all data samples. As expected, *length of stay* in the ward and in the hospital have the highest impact on the predictions. The Shapley analysis results showed that the longer the stay in a ward or hospital, the more likely it is for a patient to be classified by the model as colonised. Conversely, the shorter the stay in a ward or the hospital, the more likely to be classified as non-colonised. The number of patients in a ward also has an important impact on model predictions. The higher the number of patients in the ward, the more probable the model output to be positive (colonised). Despite its lower impact, *female* gender influenced the model output in the positive (colonised) direction compared to *male*, which has the opposite effect. This could be explained by the fact that that most prevalent bacteria in the dataset were *E. coli* and that urinary tract infections are more common among women than men^47^. Similarly, the *neonatal intensive care unit* (NICU) was less important to the model decisions than the *medical intensive care unit* (MICU) and *surgical intensive care unit* (SICU). A patient in SICU and MICU will more likely drive the model towards a positive output (colonised), while a patient in NICU will more likely drive the model towards a negative output (non-colonised). These findings are aligned with previous risk factor analysis studies for nosocomial infections in adult intensive-care units^48^.

**Figure 6:**
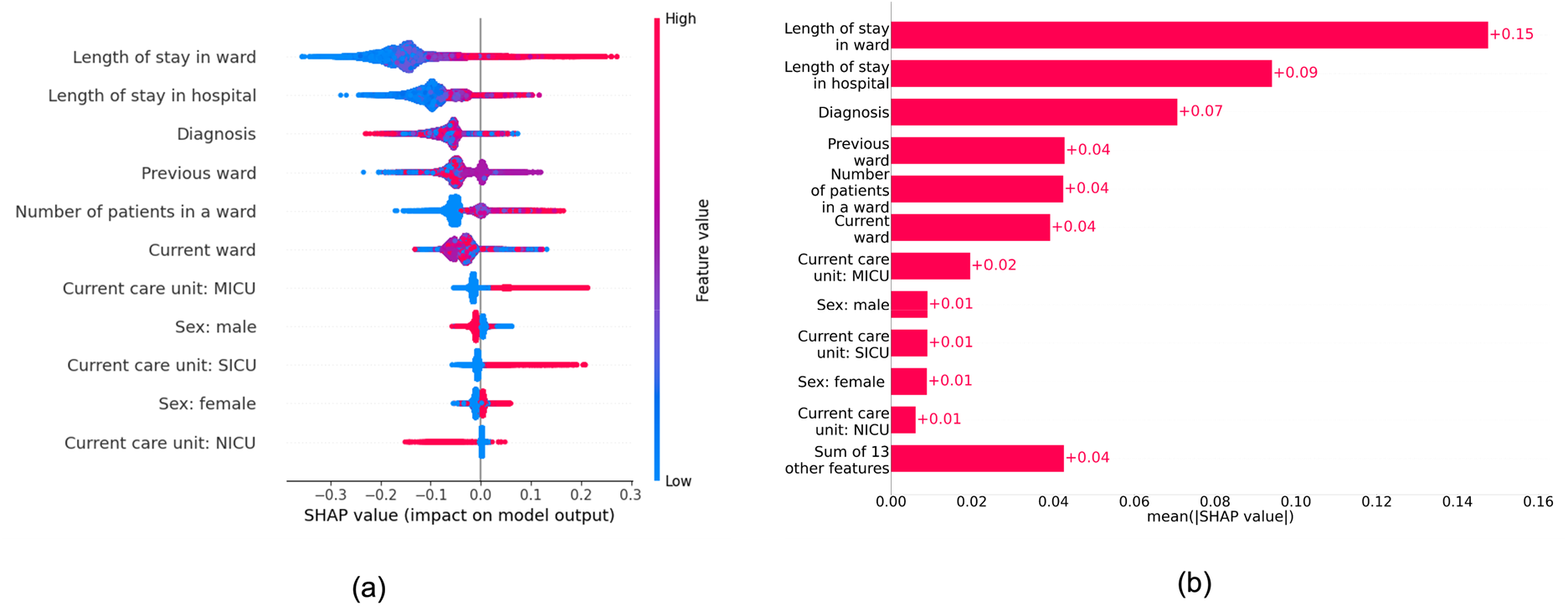
Feature contribution to colonisation risk prediction. a) Shapley values for top-11 features, sorted by their impact on model predictions. b) Mean absolute value of every feature presented in a).

### Colonisation path analysis

A major advantage of using graph models and GNNs to predict colonisation risks is that they naturally provide possible transmission paths via graph edges. In Figure 7, we show three examples of patients that were classified correctly as colonised by the *GNN - all* model: nodes 57627 (top left), 154208 (top right) and 211904 (bottom). Nodes in green represent non-colonised patients and nodes in red represent positive culture for Enterobacteriaceae. Filled colours represent colonised patients. In the scenario of Figure 7 - top left, patient 57627 (focus patient hereafter), who was colonised by *K. pneumoniae*, stayed in the hospital for 9 days and was directly linked to four patients: two in the same room (one non-colonised and one colonised) and two in different rooms (both non-colonised). Similar to the focus patient, patient 119123 was colonised by *K. pneumoniae* and had the longest hospital stay in this subnetwork (11 days). Thus, if both bacteria strains were genetically identical (or derived phylogenetically), a possible transmission route could have been from patient 119123 to the focus patient or vice-versa, or from a common source within the ward (e.g., door handle). In Figure 7 - top right, patient 154208 (focus patient hereafter) stayed for 24 days in the hospital and had an immediate link to patient 23117 (non-colonised) from a different ward via a healthcare worker, and a second-degree connection to patient 111558 (colonised) from another ward. The latter patient and the focus patient were both colonised by *K. pneumoniae*, like in the previous scenario. Hence, the path 111558-23117-154208 could be one of the possible transmission routes within the hospital. For the third scenario, Figure 7 - bottom, patient 211904 (focus patient hereafter), male, stayed for 10 days and had a direct connection to patient 36255 via the same ward, both colonised, but by different bacteria. Moreover, these patients had a second-degree connection to patient 158476, female, via a healthcare worker link, who was colonised by *E. coli*, as the focus patient. Since patient 158476 was hospitalised for 7 days, she may have been colonised by the same strain as the one of the focus patient (or vice-versa), who may have been previously colonised. Thus, the undirected path 211904-36255-158476 could be a possible transmission route. Nevertheless, exact identification of transmission routes for such scenarios would require detailed phylogenetic analysis of bacterial samples^49^.

**Figure 7:**
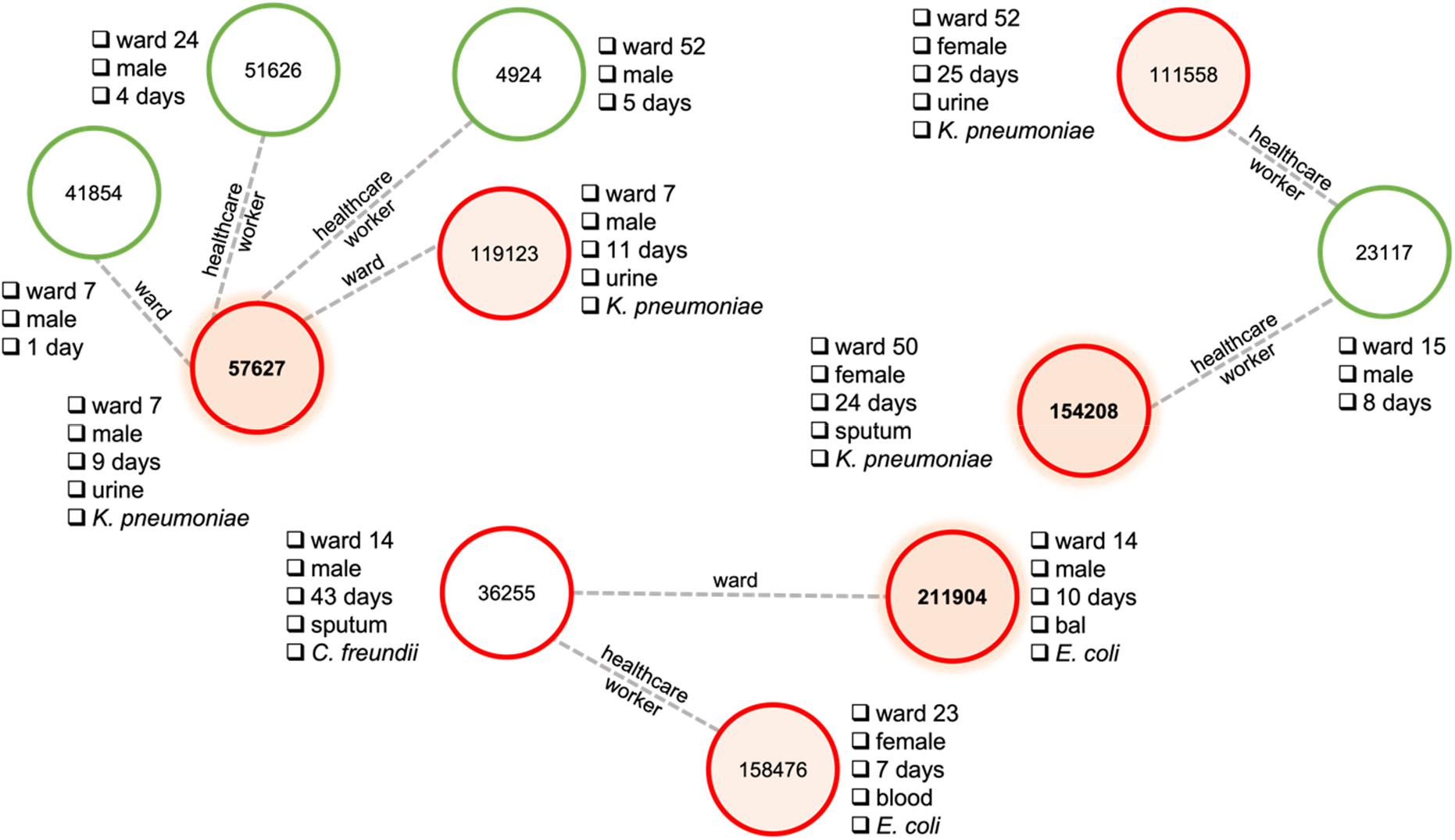
Bacteria transmission scenarios via graph paths. Green nodes: non-colonised patients; red nodes: colonised patients.

## Discussion

This study describes a machine learning model based on graph neural networks to predict patients at risk of colonisation by AMR and MDR Enterobacteriaceae. We model the data as a graph to represent possible connections and interactions between patients and healthcare workers inside the healthcare facility. Different graph topologies were proposed based on geographic location and interaction with healthcare workers. We considered spatiotemporal features, such as length of stay and ward movement, in addition to clinical and laboratory information, to encode patients via node features in different graph topologies. Performance analyses showed that GNN models provide robust predictive performance, often above AUROC of 92%, outperforming all classic machine learning baselines used in our experiments. These results demonstrate the importance of incorporating topological features to learn patterns of patient profiles that are more likely to be colonised by MDR Enterobacteriaceae.

Other recent studies investigated the use of machine learning to predict colonisation risk of AMR species of the Enterobacteriaceae^27,28^, Enterococcaceae^26^ and Staphylococcaceae^18^ families, achieving robust predictive performance with an AUROC between 88% and 89%. Our study is the first to consider the colonisation risk for AMR and MDR Enterobacteriaceae family, which are responsible for the highest incidence of nosocomial infections and HAI-related mortality^50^, using a transmission network approach and spatiotemporal information. Moreover, in contrast to previous studies, which were based on ensemble of tree methods such as random forest, our proposed methodology used a deep learning approach and showed superior predictive power for the colonisation prediction problem of Enterobacteriaceae (in our experiments, 86% for random forest vs. 93% for GNN). Another advantage of the graph-based modelling, as opposed to tabular data used in previous studies, is that possible transmission routes can be inherently extracted from the model, opening an avenue for data-driven transmission route hypothesis generation.

Following IPC guidelines, when an AMR Enterobacteriaceae outbreak occurs in a hospital or in a long-term care facility, colonised patients are initially isolated. Then, the contact group, i.e., patients potentially colonised by the outbreak strain, is identified to determine the magnitude of the outbreak and, if required, additional IPC measures are applied^51^. Using administrative information from the EHR system, contact tracing information can be obtained and used to determine other patients potentially at risk, which will ultimately go through a screening process to duly confirm colonisation by the AMR strain. This process is reactive and can be uncomfortable for patients, as well as very costly and time consuming, preventing thus corrective actions to be taken in due time^52^. The predictive model proposed in this study could help improve IPC measures against Enterobacteriaceae, and other pathogens, in several ways. First, it could help to estimate the contact group with high accuracy, which in turn could lead to more effective measures to curb transmission and infection. Second, possible transmission paths could be automatically derived from the graph model, providing hypotheses for transmission routes. Lastly, and more importantly, if deployed in a surveillance mode, it could support early identification of potential patients at risk of AMR and MDR colonisation and enable outbreak forewarning, which could have an even bigger positive impact on live-saving and financial costs.

Despite the black-box nature of neural networks, explainable artificial intelligence methods, such as the Shapley values used to analyse our results, can provide an effective approach to interpret the model decisions and support identification of risk factors associated to colonisation risks. Among the features having the highest impact on model predictions, features such as *length of stay, previous ward* and *gender* have also been identified as relevant by previous epidemiological studies that investigated risk factors for HAI colonisation and infection. For example, Patel *et al*.^53^ showed that carbapenem-resistant *K. pneumoniae* infection was independently associated with longer length of stay before infection. McHaney-Lindstrom *et al*.^54^ showed that unit transfer increases the odds of contracting an infection by 7%. For the case of gender, the model not only identified this feature as a risk factor but also showed that being a female is associated with higher risk of Enterobacteriaceae colonisation. This result was found in previous risk analysis studies, which identified higher incidence rates of *E. coli* in females as compared to males^55^.

Applying machine learning algorithms to solve the task of colonisation risk prediction is challenging due to the imbalanced nature of the data. Machine learning models are often biased towards the majority class (i.e., non-colonised in our case), and in the worst-case scenario, they will ignore the minority group entirely. In such cases, accuracy and other micro-average metrics are not optimal to evaluate model performance. Even if the model fails to predict the minority class, i.e., *colonised* in our case, accuracy might still be high due to the high percentage of non-colonised patients. To provide a more comprehensive view of our results, we reported macro-averaged metrics, which assign equal weights to positive (colonised) and negative (non-colonised) classes. Moreover, we reported these same metrics in a balanced scenario, using an undersampling technique^56^. In both cases, results showed that the models learned colonisation patterns, with macro F1-score well above the 50% threshold indicating that some colonisation patterns were indeed learned by the model.

Our study has several limitations, both in terms of data and modelling. First, the model might not be able to generalise to other hospitals as it was only evaluated in a single hospital unit dataset. Indeed, it is known that the epidemiology of HAI varies within different units and geographies^57^. Investigations of generalization performance for this type of models will warrant specific future research. Second, while we avoided using predictors that might overlap with the dependent variable, such as antimicrobial consumption (e.g., trimethoprim-sulfamethoxazole antimicrobial medication could be a predictor for *E. coli* infection^58^), other predictors, such as diagnosis at admission, could still have caused prediction bias. Nevertheless, given the distribution of diagnoses in the dataset, we expect that this bias is limited, if any. Third, our graph topology does not include environmental transmission, while it is known that indirect transmission via the environment is an important part of HAI routes^59^. Due to the lack of fine-grained contact and sampling data in the MIMIC-III dataset, environment-related transmission pathways were ignored in our models as this scenario could not be realistically captured. Understanding the impact of environmental transmission on model performance could be another research direction. Lastly, due to the anonymisation strategy of MIMIC-III and, more specifically, to the time shift, the data used in our experiments could be better regarded as a synthetic data (generated from real data) rather than as real hospital data^60,61^.

To conclude, this study shows that encoding topological information about patient-healthcare worker interactions using GNNs can improve predictive performance of AMR/MDR Enterobacteriaceae colonisation models and support identification of patients potentially at risk of infection. Hence, these models could be used to enhance IPC programmes and reduce HAI burden. Given the data-driven approach of our method, we expect that it could be expanded to other pathogens with similar transmission dynamics and to other healthcare settings.

## Data Availability

The data that support the findings of this study are available from the MIMIC-III database, which is provided by the MIT Lab for Computational Physiology and can be accessed at https://mimic.physionet.org. MIMIC-III is a large, freely-available database comprising de-identified health-related data associated with over forty thousand patients who stayed in critical care units of the Beth Israel Deaconess Medical Center between 2001 and 2012. Please note that access to the MIMIC-III database requires the completion of a data use agreement and proof of completion of a course in human subject research ethics, such as the CITI 'Data or Specimens Only Research' course.

## Data sharing

The MIMIC-III database is freely and publicly available through PhysioNet. The code will be shared on GitHub upon acceptance.

## Authors and contributors

RG designed and implemented the models, ran the experiments and analyses. RG and DT wrote the manuscript draft. DT and SGP conceptualised the experiments and acquired funding. RG, DP and SGP curated the data. RG, AB, DP and DT analysed the data. All authors reviewed and approved the manuscript.

